# Cognitive predictors of vaccine hesitancy and COVID-19 mitigation behaviors in a population representative sample

**DOI:** 10.1101/2022.01.02.22268629

**Authors:** Anna Hudson, Peter A. Hall, Sara C. Hitchman, Gang Meng, Geoffrey T. Fong

**Affiliations:** School of Public Health Sciences, University of Waterloo, Waterloo, Ontario, Canada; Department of Psychology, University of Waterloo, Waterloo, Ontario, Canada; Centre for Bioengineering and Biotechnology, University of Waterloo, Waterloo, Ontario, Canada; Department of Communication and Media Research, University of Zurich, Zürich, Switzerland; Ontario Institute for Cancer Research, Toronto, Ontario, Canada

**Author notes:** Equal contribution (Anna Hudson, Peter A. Hall).

**Keywords:** COVID-19, SARS-CoV-2, behavior, cognition, vaccine hesitancy

## Abstract

With the continued threat of COVID-19, predictors of vaccination hesitancy and mitigation behaviors are critical to identify. Prior studies have found that cognitive factors are associated with some COVID-19 mitigation behaviors, but few studies employ representative samples and to our knowledge no prior studies have examined cognitive predictors of vaccine hesitancy. The purpose of the present study, conducted among a large national sample of Canadian adults, was to examine associations between cognitive variables (executive function, delay discounting, and temporal orientation) and COVID-19 mitigation behaviors (vaccination, mask wearing, social distancing, and hand hygiene). Findings revealed that individuals with few executive function deficits, limited delay discounting and who adopted a generally future-orientation mindset were more likely to be double-vaccinated and to report performing COVID-19 mitigation behaviors with high consistency. The most reliable findings were for delay discounting and future orientation, with executive function deficits predicting mask wearing and hand hygiene behaviors but not distancing and vaccination. These findings identify candidate mediators and moderators for health communication messages targeting COVID-19 mitigation behaviors and vaccine hesitancy.

## 1. Introduction

Understanding the determinants of COVID-19 mitigation behaviors is critically important for managing the current pandemic and future infectious disease outbreaks. Beyond vaccination, the most widely recommended individual mitigation behaviors for COVID-19 are mask wearing, physical distancing and hand hygiene^1^. These behaviors collectively require consistent implementation in a variety of social contexts and continually changing circumstances.

Behavioral consistency may require attention to cumulative benefits to the individual and society as a whole, attention to cues that impel the behaviors, holding requirements in working memory, and the ability to flexibly alternate between implementation of behaviors (e.g., mask wearing) with dynamically changing environments (e.g., inside versus outside, in the presence of others versus alone). A mindset oriented to future benefits, strong valuation of non-immediate contingencies, and relatively intact executive functioning may be critical.

Consistent with this logic, several studies have shown that executive function is associated with adherence to mitigation behaviors in adults^2–4^ and in older populations ^5^. Similarly, greater temporal discounting and risky decision-making is associated with less mask-wearing and social distancing behavior^6,7^. Finally, future-oriented thinking has been shown to increase satisfaction and compliance with COVID-19 restrictions ^8^. However, because few studies have examined all three factors within the same sample, their comparative importance of these three factors is largely unknown.

The largest study to date examined 850 adult participants from the United States, available via Amazon Mechanical Turk, in the opening weeks of the pandemic (i.e., before vaccines were available, and many mitigation behaviors were not yet mandated). Findings revealed that one subcomponent of executive function (working memory) predicted social distancing compliance^3^. A more comprehensive examination of cognitive predictors and behavioral outcomes is largely absent, and there are no other studies examining cognitive predictors of vaccination status specifically. Some studies involving prediction of vaccination status are limited by a relatively low proportion of vaccine hesitant participants; this is most likely to occur in countries wherein vaccine uptake is high. This limitation can be rectified by quota sampling to increase the proportion of the overall sample that is vaccine hesitant relative to vaccinated.

The current study was intended to examine cognitive predictors of COVID-19 mitigation behaviors and vaccine hesitancy in a nationally representative sample of Canadian adults between the ages of 18-55. At the time of the data collection (November, 2021), mandatory indoor mask wearing was mandated in most provinces, and recommendations for hand hygiene and distancing were well known and reminders ubiquitous; vaccines were widely available for all adults and strongly recommended country-wide. Within this context, it was hypothesized that stronger executive function, lower discounting of future rewards, and a more future-oriented mindset would be associated with higher vaccination rates and more consistent performance of COVID-19 mitigation behaviors. This study advances existing literature by examining three conceptually important cognitive and motivational variables using a large population-based dataset with sampling and statistical methods that allows for generalization to the population.

## 2. Methods

### Participants

Participants were respondents in Wave 1 of the Canadian COVID-19 Experiences Survey (CCES ^9^), a national cohort survey of 2002 Canadian adults aged 18-55 (Mean = 37, *SD* = 10.4; 60.8% female). The cohort was recruited to have an equal proportion of vaccinated and vaccine hesitant individuals. In the recruited sample, 50.2% reported receiving two vaccine shots (i.e., fully vaccinated by the standards at the time of data collection), and 43.3% reported receiving no vaccinations. Further 5.5% reported receiving one vaccine shot but were not intending to receive a second shot.

### Procedure

The online survey was administered between September 28^th^ and October 21^st^, 2021. Participants were contacted by email with an invitation to participate in the survey, with a link provided to all eligible participants. A quota target of equal numbers of vaccinated and vaccine hesitate was applied to ensure an equal sample of vaccinated and vaccine hesitant individuals. Within each quota target, participants were recruited across ten Canadian provinces. The survey firm (Leger) and the University of Waterloo research team monitored the survey to ensure the final sample reached the intended quota targets. The authors assert that all procedures contributing to this work comply with the ethical standards of the relevant national and institutional committees on human experimentation and with the Helsinki Declaration of 1975, as revised in 2008. Informed consent was obtained from all participants in this study.

### Measures

#### Executive dysfunction

Symptoms of executive dysfunction were assessed across four ‘self-restraint’ subscale items from the Barkley Deficits in Executive Functioning Scale short-form (BDEFS-SF). The following four items were used: “I am unable to inhibit my reactions or responses to events or to other people”, “I make impulsive comments to others”, “I am likely to do things without considering the consequences for doing them”, and “I act without thinking”. Participants were asked to report how often they have experienced each of the four problems over the past 6 months on a numeric scale, where 1=“never or rarely”, 2=“sometimes”, 3=“often”, and 4=“very often”. The four items were z-transformed and averaged together to create a composite executive dysfunction measure, with higher scores reflecting more dysfunction. Because values were positively skewed, a Log10 transformation was applied. Cronbach’s alpha for the 4-item scale was 0.815, indicating good reliability.

#### Delay Discounting

A validated 5-item delay discounting (DD) task was used to assess valuation of non-immediate contingencies. The 5-item DD task presents respondents with a series of choices between a fixed immediate monetary amount ($500) and a larger reward at varying delay times (i.e., “Would you rather have $500 now, or $1000 in 4 hours; 1 day; 3 weeks; 2 years?” ^10^). From these an indifference point can be calculated, reflecting the time at which the preference for a larger later reward reverts to a preference for the smaller immediate reward. This value is denoted by k. Higher *k* values are indicative of more impulsive decisions, preferring a lower immediate reward over waiting for a higher reward, that is, a higher discount rate. Because *k* values were positively skewed, a Log10 transformation was applied. Because k values are between 0 and 1, log k values are negative, and lower log k values are associated with greater impulsivity.

#### Time Perspective

Participants responded to 4 questions assessing their degree of present and future orientation. Participants responded from 1=“strongly agree” to 5=“strongly disagree”, with 3=“neither agree nor disagree” to two present-orientation questions (i.e., “Living for the moment is more important than planning for the future”, and “A spend a lot more time thinking about today than thinking about the future”) and to two future-orientation questions (i.e., “I spend a lot of time thinking about how my present actions will have an impact on my life later on”, and “I consider the long-term consequences of an action before I do it” ^11^. Participants responding with “Refused”, or “Don’t know” were removed from analyses (*n*=179). The two present perspective, and two future perspective questions were first standardized and averaged together to form separate subscales for present and future orientation. The two subscales were subtracted such that higher scores represented greater future relative to present orientation. Cronbach alphas for each of the subscales indicated acceptable reliability (present orientation: α=0.742; future orientation: α=0.665)

#### Mitigating Behaviors

Participants responded to questions assessing social distancing (“When outside your home, how consistently do you currently maintain a distance from others of at least 2 meters?”), mask wearing (“How often do you currently wear a mask when you are in INDOOR public places?”), and hand hygiene (“How often do you thoroughly wash your hands during the day?”). Participants responded using the following response options: 1=“Not at all”, 2= “Rarely”, 3= “Sometimes”, 4= “Most of the time” and 5=“All of the time”. Higher scores on these items reflected an increased consistency in behavioral performance. Participants responding “Refused”, or “Don’t know” to the items were removed from analyses (*n*=74). The social distancing item module also contained a “I haven’t had contact with others” response, and the mask wearing item module contained a “I am never in indoor public places” response. Participants giving these responses (*n*=49) were also removed, as it was assumed that such participants did not have an opportunity to enact the response being queried (e.g., immunocompromised individuals avoiding all indoor public spaces).

#### Vaccination status

Vaccination status was queried using the following item: “Have you received any COVID-19 vaccine shots?” Responses available were as follows, “I have NOT received any vaccine shot”, “Received ONE vaccine shot”, “Received TWO vaccine shots” [coded as fully vaccinated], or refused/don’t know. Those indicating that they had received only one shot were asked the following additional question: “What best describes your intention to get your next shot?” Response options were as follows: “I have NO plan to get a second shot”, “I am unsure whether I will get the second shot” [coded as unvaccinated without intention], and “I plan to get the second shot, but have NOT yet scheduled an appointment”, and “I am planning to get the second shot and have scheduled an appointment.”

#### Demographics

Age, sex and income assessed by respondent report. Geographical region was coded directly from the online survey profile of each respondent.

### Statistical Analysis

All analyses were conducted in SPSS version 27.0.1.0. Separate hierarchical multiple regression models were conducted, predicting behavioral outcomes from the following predictors: (1) BDEFS score, (2) Delay Discounting (k-value), (3) TPQ score (future orientation). Control variables were entered on the first step, followed by main effects and interactions on subsequent steps. As such, all analyses were examined while controlling for demographic factors and vaccination status moderation effects were tested. Further sensitivity analyses were performed wherein income, education and geographic region (province) were added as covariates. From the *p*-values for each focal predictive analysis, heat maps were constructed and displayed in original form, and with *p*-values corrected for false discovery rate (FDR).

## 3. Results

### Vaccination Status

With reference to fully vaccinated, Higher BDEFS scores did not predict significantly increased odds of being unvaccinated (*OR*=0.94, 95% *CI* [0.78-1.13], *p*=.492), but did predict higher likelihood of being partially vaccinated without intention to be fully vaccinated (*OR*=1.90, 95% *CI* [1.38-2.61], *p*<.001). Those showing higher impulsivity on the delay discounting task were more likely to be unvaccinated (*OR*=1.24, 95% *CI* [1.11-1.39], *p*<.001) and partially vaccinated without intentions to be fully vaccinated (*OR*=1.43, 95% *CI* [1.15-1.78], *p*=.001). Similarly, greater future orientation predicted lower odds of being unvaccinated (*OR*= 0.82, 95% *CI* [0.69 -0.99], *p*=.034) and lower odds of being partially vaccinated without intention to be fully vaccinated (*OR*=0.56, 95% *CI* [0.39-0.82], *p*=.003). Figure 1 presents a heat map showing *p* values associated with cognitive measures as predictors of vaccination status (Table 1).

**Figure 1.**
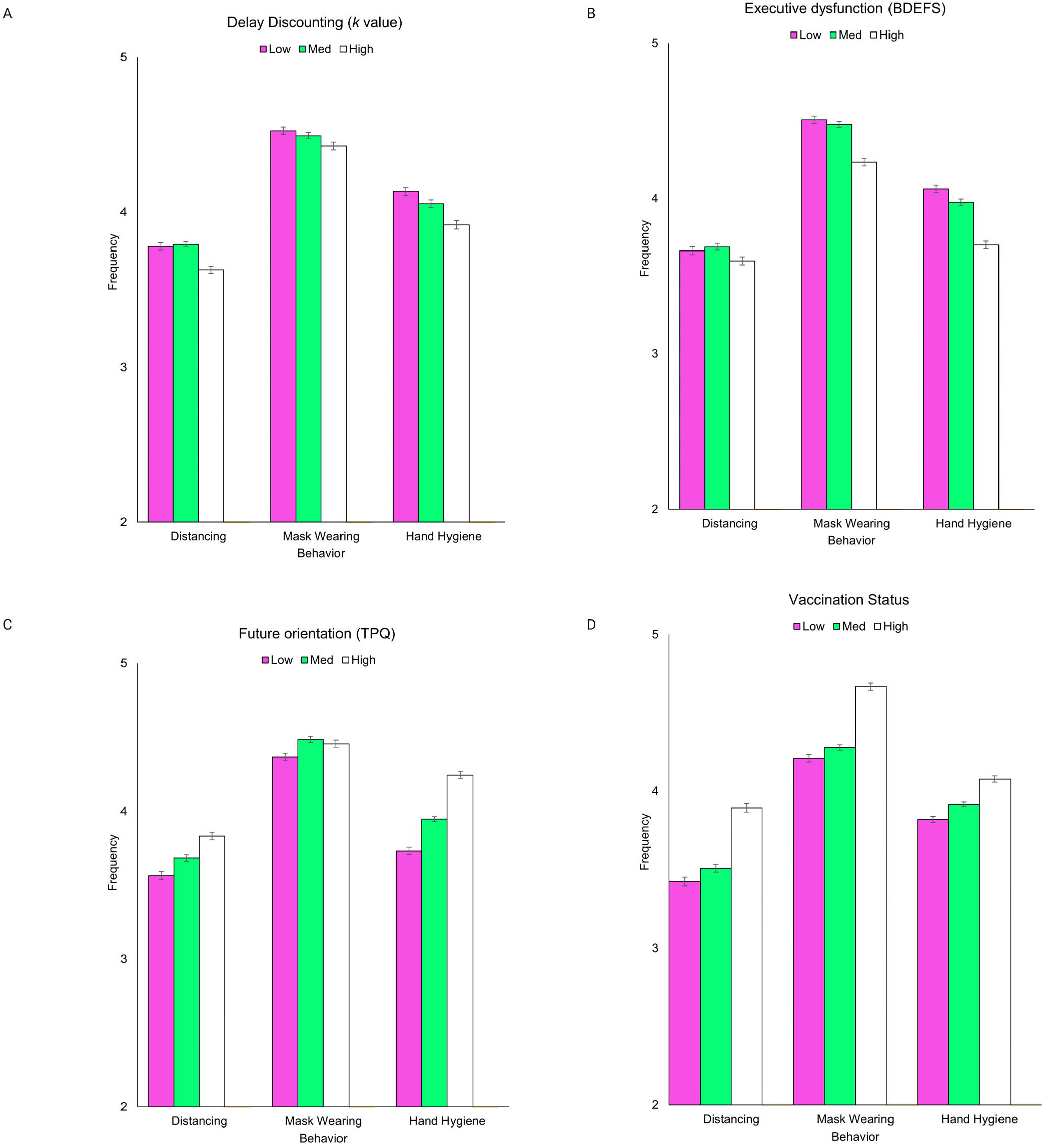
Main effects of a) delay discounting, b) executive dysfunction, c) future orientation and d) vaccination status on frequency of COVID-19 mitigation behaviours. Higher scores on the y-axis reflect increased frequency of behavior performance. Participants were split into the lower (pink), mid (green), and higher categories (black) based on z-scores (−1.0, 0, +1.0). Error bars represent standard errors. Created with BioRender.com

**Table 1.** Heat map of p-values for cognitive measure crossed with mitigation behaviors and vaccination status.

|  | Masks | Social<br>Distancing | Hand-<br>hygiene | Vaccination<br>status | no intention<br>for 2<sup>nd</sup> shot |
| --- | --- | --- | --- | --- | --- |
| BDEFS | <.001 | .226 | <.001 | .492 | <.001 |
| DD | .046 | .051 | .086 | <.001 | .001 |
| TP | .014 | .012 | .005 | .034 | .003 |
| FDR-corrected values |  |  |  |  |  |
| BDEFS | .003 | .242 | .008 | .492 | .003 |
| DD | .063 | .064 | .099 | .003 | .003 |
| TP | .023 | .012 | .011 | .034 | .008 |

Note: Green cells indicate statistically significant effects; yellow cells indicate marginal effects and red/orange cells indicate non-significant effects. BDEFS=Barkley Deficits in Executive Function Scale; DD=Delay discounting (k value); TP=time perspective (future orientation). FDR=false discovery rate, calculated as per Benjamini and Hochberg <sup>[12]</sup>.

### Mitigation Behaviors

Higher BDEFS scores were associated with lower frequency of mask wearing (β= -0.133, 95% *CI* [-0.162, -0.082] *p*<0.001) and hand hygiene behaviors (β= -0.139, 95% *CI* [-0.184, -0.095], *p*<0.001; Figure 1). BDEFS score was not significantly associated with the frequency of social distancing behaviors (β= -0.027, 95% *CI* [-0.082, 0.019], *p*=0.226; Table 2; Figure 1). Greater delay discounting (higher *k* scores) were marginally associated with more consistent social distancing behaviors (β=0.044, 95% CI [0.000, 0.101], *p*=0.051), mask wearing (β=0.044, 95% CI [0.001, 0.081], *p*=0.046) and hand hygiene behaviors (β=0.039, 95% CI [-0.005, 0.084], *p*=0.086). Greater future orientation was associated with more consistent mask wearing (β=0.054, 95% CI [0.010, 0.092], p=0.014), hand hygiene (β= 0.064, 95% CI [0.023, 0.129], *p*=0.005), and compliance with social distancing behaviors (β=0.056, 95% CI [0.015, 0.117], *p*=0.012).

**Table 2.** Regression analyses predicting mitigation behaviors.

| **<u>Variables</u>** |  | **<u>Social Distancing</u>** |  | **<u>Mask Wearing</u>** |  | **<u>Hand Hygiene</u>** |  |
| --- | --- | --- | --- | --- | --- | --- | --- |
|  |  | Beta<br>[95% CI] | *p* | Beta<br>[95% CI] | *p* | Beta<br>[95% CI] | *p* |
| **Executive**<br>**function**<br>**(Mean)** | BDEFS | -0.027<br>[-0.082, 0.019] | 0.226 | -0.133<br>[-0.162, -0.082] | <0.001 | -0.139<br>[-0.184, -0.095] | <0.001 |
|  | Vaccination | 0.186<br>[0.163, 0.265] | <0.001 | 0.239<br>[0.183, 0.263] | <0.001 | 0.111<br>[0.067, 0.155] | <0.001 |
|  | BDEFS*Vaccination | -0.028<br>[-0.085, 0.018] | 0.201 | -0.039<br>[-0.077, 0.004] | 0.076 | -0.009<br>[-0.054, 0.036] | 0.689 |
| **Delay**<br>**Discounting (DD)** | DD | 0.044<br>[0.000, 0.101] | 0.051 | 0.044<br>[0.001, 0.081] | 0.046 | 0.039<br>[-0.005, 0.084] | 0.086 |
|  | Vaccination | 0.187<br>[0.165, 0.267] | <0.001 | 0.238<br>[0.178, 0.259] | <0.001 | 0.107<br>[0.062, 0.153] | <0.001 |
|  | DD*Vaccination | -0.009<br>[-0.061, 0.040] | 0.687 | -0.010<br>[-0.049, 0.031] | 0.656 | 0.031<br>[-0.013, 0.077] | 0.166 |
| **Temporal**<br>**Orientation** | Time | 0.056<br>[0.015, 0.117] | 0.012 | 0.054<br>[0.010, 0.092] | 0.014 | 0.064<br>[0.023, 0.129] | 0.005 |
|  | Vaccination | 0.192<br>[0.174, 0.277] | <0.001 | 0.227<br>[0.172, 0.254] | <0.001 | 0.037<br>[0.023, 0.129] | 0.005 |
|  | Time*Vaccination | 0.006<br>[-0.043, 0.058] | 0.771 | 0.040<br>[-0.003, 0.077] | 0.077 | -0.034<br>[-0.093, 0.012] | 0.128 |

Note: Main effects and two-way interactions for focal predictors and vaccination status controlling for age and gender. Those reporting 'don't know'; 'refused'; and 'NA' vaccination status were classified as unvaccinated. All coefficients are standardized Beta weights.

### Main Effects and Interactions involving Vaccination Status

In all models, being vaccinated predicted stronger compliance with mitigation measures, for example in models involving BDEFS, higher frequency of mask wearing (β= 0.239, 95% CI [0.180, 0.259], *p*<0.001), hand hygiene (β= 0.111, 95% CI [0.067, 0.155], *p*<0.001) and social distancing (β= 0.186, 95% CI [0.163, 0.265], *p*<0.001). No interactions involving vaccination status were evident for BDEFS score (mask wearing: *F*(1,1954)= 3.15, *p*=0.076; hand hygiene: *F*(1,1954)= 0.160, *p*=0.689; social distancing: *F*(1,1954)= 1.63, *p*=0.201), delay discounting (mask wearing: *F*(1,1910)= 0.199, *p*=0.656; hand hygiene: *F*(1, 1910)= 1.92, *p*=0.166; social distancing: *F*(1, 1910)= 0.163, *p*=0.687), or temporal orientation (mask wearing: *F*(1, 1949)= 3.23, *p*=0.072; hand hygiene: *F*(1, 1949)=2.32, *p*=0.128; social distancing: *F*(1, 1949) = 0.085, *p*=0.771).

### Sensitivity Analysis

Further adjustment for education, income, and geographic location (coded as province) had no overall effect on the findings. In these fully adjusted models, mask wearing, hand hygiene and social distancing continued to be predictable as a function of executive dysfunction (mask wearing β= -0.129, 95% CI [-0.158, -0.079] *p*<0.001; hand hygiene β= -0.137, 95% CI [-0.182, -0.093], *p*<0.001; social distancing β= -0.027, 95% CI [-0.082, 0.019], *p*=0.225), delay discounting, (mask wearing β=0.041, 95% CI [-0.002, 0.077], *p*=0.064; hand hygiene β=0.039, 95% CI [-0.005, 0.084], *p*=0.084; social distancing β=0.043, 95% CI [0.000, 0.100], *p*=0.052), and temporal orientation (mask wearing β=.075, 95% CI [0.029, 0.111], *p*<0.001; hand hygiene β=.071, 95% CI [0.031, 0.137], *p*=0.002; social distancing β=0.064, 95% CI [0.024, 0.127], *p*=0.004).

## 4. Discussion

In this population-based representative sample, we examined cognitive determinants of COVID-19 vaccination and mitigating behaviors among adults between the ages of 18 and 55 years. Findings demonstrated that those who possessed higher executive function, lower delay discounting, and greater future orientation were more likely to be vaccinated and engage in key COVID-19 mitigation behaviors (i.e., social distancing, mask wearing, and hand hygiene). This was true across standard demographic variables such as age and sex, and proved robust through a variety of sensitivity analyses. Among the three cognitive variables, delay discounting and future orientation were the most consistently predictive of COVID-19 vaccination and mitigation behaviors.

This pattern suggests that value processing wherein non-immediate outcomes are protected from discounting is generally important in COVID-19 mitigation, as is a conscious appreciation of (and belief in) connections between present actions and later outcomes. Although there are links to neurobiological substrates, delay discounting and future orientation are potentially malleable cognitive processes, and public health communications directing attention to the value of non-immediate outcomes may be particularly important to deploy population wide. Executive dysfunction, on the other hand, was associated with a more circumscribed set of COVID-19 mitigating behaviors. Specifically, higher executive dysfunction predicted less consistent mask wearing and hand hygiene—both behaviors are repetitive, discrete behaviors and acts of commission. Measurement considerations are potentially important as well: the BDEFS is a deficit-based assessment tool designed for clinical practice, and it may therefore miss some important dimensional aspects of cognitive function more relevant to vaccination and distancing in a population survey format. Notably, the latter two outcomes are also highly multi-determined, such that political orientation and other beliefs may overshadow the predictive power of any relatively coarse cognitive indicator.

Among the three mitigation behaviors examined, social distancing was least predictable from the neurocognitive variables tested. It is possible that social distancing can be better predicted from social-cognitive variables (e.g., beliefs about and attitudes towards other individuals and groups), given its links to the social fabric of everyday life, as opposed to the physical environment and de-contextualized behavior. Given that social distancing is fundamentally a relational behavior with interpersonal consequences, this possibility may warrant further investigation.

Our findings are consistent with several other studies that have found associations between executive functions and mitigation behaviors^2–5^. However, our study extends these prior studies via inclusion of delay discounting, and our findings that the associations are no different across vaccinated and vaccine hesitant individuals. The latter cannot be tested with sufficient power in samples that do not contain a high proportion of both vaccinated and vaccine hesitant individuals. Our study—which used a sampling quota such that a large and approximately equal numbers of fully vaccinated and vaccine hesitant individuals were surveyed—is the largest study conducted to date, allowing for a strong test of the hesitancy moderation hypothesis. Likewise, although there are reliable effects of vaccination status on implementation of mitigation behaviors, such effects are largely independent from the effects of cognitive variables on the implementation of mitigation behaviors.

Strengths of the current investigation include the use of a population representative sample, ensuring the findings may be generalized to the larger population from which they were drawn. Additionally, the use of quota sampling to ensure approximately 50% vaccine hesitant ensures adequate statistical power to determine the moderating impact of vaccination status on any findings. No other investigations to date have this feature and would for the most part be unable to determine uniformity of prediction across vaccination status groups. Limitations include the use of self-reported vaccination status and executive function, and abbreviated versions of time perspective measures due to the population survey format. Finally, the sample age range is from 18-55 years, thereby excluding older adults and adolescents. However, this working age population is arguably a key population in which to study mitigation behaviors, as such individuals tend to be highly mobile and more variable in implementation of precautions than older age groups.

## 5. Conclusion

In conclusion, our findings suggest that cognitive variables reflecting future-oriented thinking, evaluative processing and behavioral control are associated with likelihood of being fully vaccinated against COVID-19 and predict more consistent implementation of mitigation behaviors. Among the three constructs, delay discounting and future orientation were the most consistent predictors of COVID-19 mitigation behaviors and vaccination status. Health communication campaigns that reinforce and emphasize positive valuation of future outcomes, and connections between present actions and later outcomes, may facilitate better response from the general public. However, it is also possible that among the less observant public, behavior may be more influenced by communications emphasizing more immediate benefit.

## Data Availability

Data will be available upon reasonable request to either of the corresponding authors.

## Data Availability Statement

Data will be available upon reasonable request to either of the corresponding authors.

## Research ethics statement

This study protocol was reviewed by and received approval from the University of Waterloo Office of Research Ethics.

## Acknowledgements

We thank Anne C.K. Quah and Thomas Agar for their assistance with survey design and management.

